# Efficacy of the PragmaVAC Manual Negative Pressure Wound Therapy Device to Treat Acute Traumatic Wounds in a Conflict Setting: A Retrospective Cohort Study from Gaza

**DOI:** 10.64898/2026.06.04.26354740

**Authors:** Ibrahim Ramadan, Mahmoud Hariri, Omar Shalakhti, Jude Alawa, Amandine Godier-Furenemont, Abd Al-Rahman Traboulsi, Hani Mowafi

## Abstract

**Background:** Acute war-related traumatic wounds present significant challenges due to significant soft-tissue damage/loss, risk of contamination, limited access to antimicrobial therapy, need for delayed closure, and limited access to surgical and wound care. Negative Pressure Wound Therapy (NPWT) has been used effectively to reduce the volume of soft-tissue defects, edema, and infection in traumatic wounds, and to promote growth of healthy granulation tissue. However, conventional NPWT devices are costly and electricity-dependent, limiting their utility in conflict settings.

**Methods:** This retrospective cohort study evaluated the use of *PragmaVAC*, a manually operated, electricity-independent NPWT device, in patients across three hospitals in Gaza with conflict-related wounds that were deemed by the treating surgeon to be unsuitable for primary closure. Secondary analysis was performed of clinical records of patients treated with the PragmaVac NPWT device to assess ability to achieve a primary outcome of wound bed with healthy granulation tissue, time to primary outcome, and rates of adverse effects. Secondary outcome of wound closure and closure method was also assessed.

**Results:** Treatment with PragmaVAC manual NPWT was prescribed to 88 patients. Of those, 27 (30.7%) had incomplete documentation of their wound healing or were lost to follow up. The remaining 61 (69.3%) had complete documentation of their wound healing, complications, and outcome with 59 (67%) successful closure and 2(2.3%) failure.

**Conclusion:** The use of the PragmaVAC NPWT device provided a safe, effective wound care option to achieve wound closure for large conflict-related traumatic wounds in resource-limited settings. Future studies may further evaluate such use through prospective trials, evaluations of patients’ experiences with manual NPWT, and evaluating outcomes beyond primary wound closure to include medium- and long-term complications, cosmesis, and cost of therapy.

## Background

Traumatic injuries with high-energy mechanisms such as blasts, gunshots, and crush injuries account for a substantial burden of war-related surgical morbidity.^1–3^ Click or tap here to enter text. Such wounds are characterized by large volume soft-tissue damage or loss, post-traumatic edema, and risk for contamination, all of which can negatively affect ability to achieve wound closure or healing. Negative Pressure Wound Therapy (NPWT) is a cornerstone of complex wound care, applying controlled sub-atmospheric pressure to the wound bed to accelerate healing. Mechanistically, NPWT promotes granulation tissue formation, reduces edema, clears exudate and bacteria, and enhances local perfusion.^4–7^ Clinical studies and meta-analyses report that NPWT can increase wound closure reduce hospital stay by 2-5 days, and lower infection risk from 40-50% compared to standard moist wound.^7–9^ Such benefits are particularly important in high-contamination of war-related trauma wounds, where minimizing complications and facilitating closure are critical. Despite these potential benefits, the use of conventional NPWT in conflict settings remains limited due to high cost, reliance on electricity, and logistical challenges related to equipment and ￼consumables.^10,11^

Over the past two decades, repeated episodes of armed conflict in Gaza have generated large numbers of trauma presentations characterized predominantly by blast-related and firearm injuries, resulting in extensive tissue destruction and long-term disability.^12,13^ During the most recent episode of increased conflict this pattern was again observed with very high numbers of trauma presentations from high-energy mechanisms such as explosive fragments, gunshot wounds from military grade weapons, and severe crush wounds resulting from building collapse, with over 70% requiring surgical wound management^14,15^ This high demand for surgical treatment took place at a time when hospitals themselves were bombed^16^ and those remaining operational did so under severe logistical constraints with reported shortages of anesthesia, antiseptics, sterile dressings, and clean water, all of which affected the timing and quality of surgical interventions.^17^. Intermittent power outages disrupted the use of sterilization equipment and oxygen delivery systems, complicating infection prevention and postoperative monitoring^17^Wound culture data collected during this period showed a high prevalence of multidrug-resistant organisms, highlighting the challenges of antimicrobial stewardship in settings where infection control measures are limited.^12,17^ In such environments, wound care solutions must be both clinically effective and feasible within constrained resource conditions, requiring minimal infrastructure and adaptability to field-based care settings.

PragmaVAC—a manually operated, electricity-independent NPWT device—was developed as a scalable alternative for resource-limited environments. Using a manual bellows mechanism, the device delivers therapeutic negative pressure (−80 to −125 mmHg) without requiring electricity.^18^ In a previous randomized controlled trial, PragmaVAC was shown to be non-inferior to standard wound care with wet-to-dry dressings in both traumatic and atraumatic wounds, similar rates of complications, while requiring fewer dressing changes.Click or tap here to enter text. Its portability, ease of use, and minimal infrastructure requirements make it well-suited for deployment in field hospitals and crisis zones where wound care resources are scarce, electricity is limited, and wound infection risk is high.^18^ The device has been used in prior conflicts (Syria, Ukraine) but its use for traumatic wound management in conflict settings remains under-characterized.

This retrospective cohort study, using secondary analysis of clinical records from hospitals in Gaza, evaluates the clinical performance of PragmaVAC in managing traumatic wounds sustained during the Gaza war. We hypothesized that the device facilitates timely granulation tissue formation, reduces dressing burden, and maintains an acceptable safety profile, even in the context of infrastructure disruption and constrained surgical capacity. Findings aim to inform the use of manual NPWT in trauma care during humanitarian emergencies and settings where conventional solutions are not feasible.

## Methods

### Study Design

This retrospective cohort study used secondary analysis of clinical data collected from patients with acute war-related traumatic in Gaza war that were treated with the PragmaVAC manual negative pressure wound therapy (NPWT) device. The study objective was to evaluate the efficacy and safety of the PragmaVac NPWT device in a resource-limited conflict zone. The study design and reporting adhere to the STROBE guidelines for observational cohort studies.

The study population was comprised of patients presenting to one of three participating hospitals with acute war-related traumatic wounds with significant soft tissue defects, whose wounds were deemed by the treating surgeon to be unsuitable for primary closure, and who were treated with the PragmaVAC NPWT device.

### Data Source

Data was obtained from the Union of Medical Care and Relief Organizations (UOSSM-USA), a humanitarian NGO supporting trauma care in Gaza. Data provided included detailed records of trauma care provided at participating hospitals including patient demographic details, interval clinical assessments with photographs of wounds when available, recorded interventions, and outcomes.

### Sample Population

Between May 2024 and May 2025, 88 patients (61 male, 27 female) were prescribed the PragmaVac NPWT device. Of those, 61(69.3%) patients had complete records of treatment and 27 (30.7%) were lost to follow up (LTFU). Patient and wound characteristics are listed in Table 1, and treatment outcomes are listed in Table 2. Inclusion criteria for this analysis included patients of any age who presented with a) war-related traumatic wounds; b) large soft tissue defects that were deemed by their treating physician to be unsuitable for primary closure; and c) who were prescribed treatment with the PragmaVAC manual NPWT device.

**Table 1.**
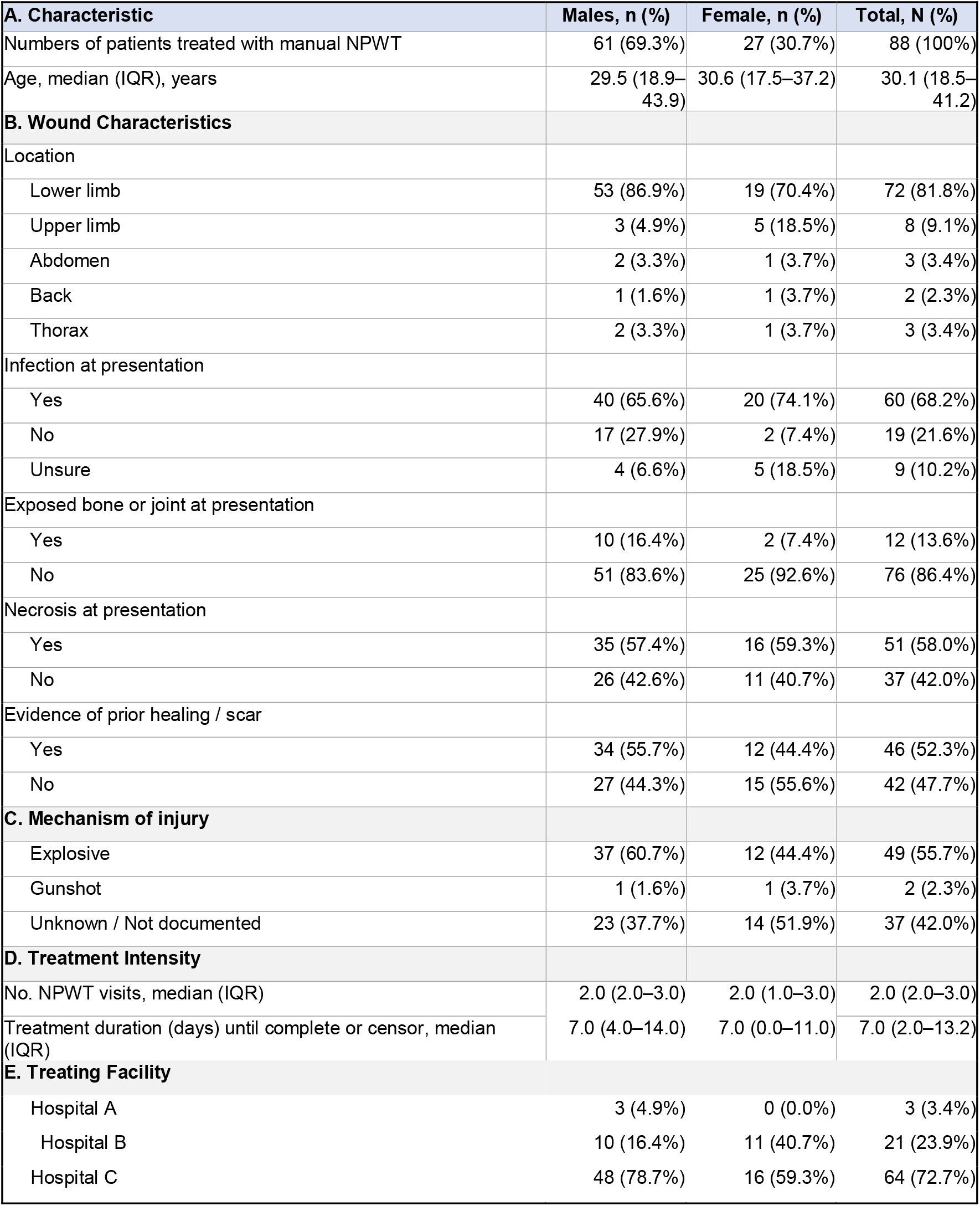
Characteristics of patients and wounds treated with manual and PWT in Gaza.

| **A. Characteristic** | **Males, n (%)** | **Female, n (%)** | **Total, N (%)** |
| --- | --- | --- | --- |
| Numbers of patients treated with manual NPWT | 61 (69.3%) | 27 (30.7%) | 88 (100%) |
| Age, median (IQR), years | 29.5 (18.9–43.9) | 30.6 (17.5–37.2) | 30.1 (18.5–41.2) |
| **B. Wound Characteristics** |  |  |  |
| Location |  |  |  |
| Lower limb | 53 (86.9%) | 19 (70.4%) | 72 (81.8%) |
| Upper limb | 3 (4.9%) | 5 (18.5%) | 8 (9.1%) |
| Abdomen | 2 (3.3%) | 1 (3.7%) | 3 (3.4%) |
| Back | 1 (1.6%) | 1 (3.7%) | 2 (2.3%) |
| Thorax | 2 (3.3%) | 1 (3.7%) | 3 (3.4%) |
| Infection at presentation |  |  |  |
| Yes | 40 (65.6%) | 20 (74.1%) | 60 (68.2%) |
| No | 17 (27.9%) | 2 (7.4%) | 19 (21.6%) |
| Unsure | 4 (6.6%) | 5 (18.5%) | 9 (10.2%) |
| Exposed bone or joint at presentation |  |  |  |
| Yes | 10 (16.4%) | 2 (7.4%) | 12 (13.6%) |
| No | 51 (83.6%) | 25 (92.6%) | 76 (86.4%) |
| Necrosis at presentation |  |  |  |
| Yes | 35 (57.4%) | 16 (59.3%) | 51 (58.0%) |
| No | 26 (42.6%) | 11 (40.7%) | 37 (42.0%) |
| Evidence of prior healing / scar |  |  |  |
| Yes | 34 (55.7%) | 12 (44.4%) | 46 (52.3%) |
| No | 27 (44.3%) | 15 (55.6%) | 42 (47.7%) |
| **C. Mechanism of injury** |  |  |  |
| Explosive | 37 (60.7%) | 12 (44.4%) | 49 (55.7%) |
| Gunshot | 1 (1.6%) | 1 (3.7%) | 2 (2.3%) |
| Unknown / Not documented | 23 (37.7%) | 14 (51.9%) | 37 (42.0%) |
| **D. Treatment Intensity** |  |  |  |
| No. NPWT visits, median (IQR) | 2.0 (2.0–3.0) | 2.0 (1.0–3.0) | 2.0 (2.0–3.0) |
| Treatment duration (days) until complete or censor, median (IQR) | 7.0 (4.0–14.0) | 7.0 (0.0–11.0) | 7.0 (2.0–13.2) |
| **E. Treating Facility** |  |  |  |
| Hospital A | 3 (4.9%) | 0 (0.0%) | 3 (3.4%) |
| Hospital B | 10 (16.4%) | 11 (40.7%) | 21 (23.9%) |
| Hospital C | 48 (78.7%) | 16 (59.3%) | 64 (72.7%) |

**Table 2:**
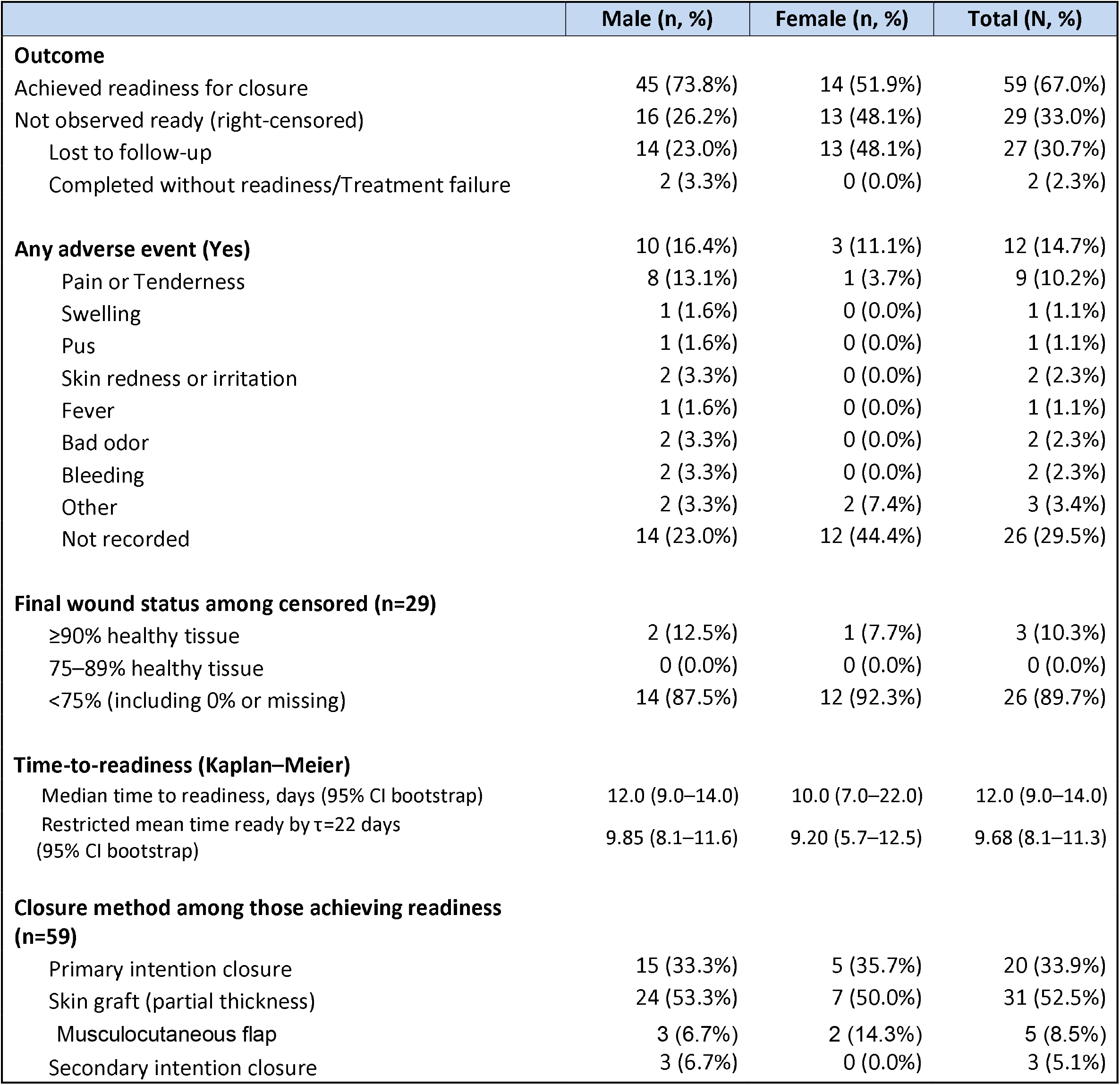
Treatment outcomes manual NPWT for war wounds in Gaza.

|  | Male (n, %) | Female (n, %) | Total (N, %) |
| --- | --- | --- | --- |
| **Outcome** |  |  |  |
| Achieved readiness for closure | 45 (73.8%) | 14 (51.9%) | 59 (67.0%) |
| Not observed ready (right-censored) | 16 (26.2%) | 13 (48.1%) | 29 (33.0%) |
| Lost to follow-up | 14 (23.0%) | 13 (48.1%) | 27 (30.7%) |
| Completed without readiness/Treatment failure | 2 (3.3%) | 0 (0.0%) | 2 (2.3%) |
| **Any adverse event (Yes)** | 10 (16.4%) | 3 (11.1%) | 12 (14.7%) |
| Pain or Tenderness | 8 (13.1%) | 1 (3.7%) | 9 (10.2%) |
| Swelling | 1 (1.6%) | 0 (0.0%) | 1 (1.1%) |
| Pus | 1 (1.6%) | 0 (0.0%) | 1 (1.1%) |
| Skin redness or irritation | 2 (3.3%) | 0 (0.0%) | 2 (2.3%) |
| Fever | 1 (1.6%) | 0 (0.0%) | 1 (1.1%) |
| Bad odor | 2 (3.3%) | 0 (0.0%) | 2 (2.3%) |
| Bleeding | 2 (3.3%) | 0 (0.0%) | 2 (2.3%) |
| Other | 2 (3.3%) | 2 (7.4%) | 3 (3.4%) |
| Not recorded | 14 (23.0%) | 12 (44.4%) | 26 (29.5%) |
| **Final wound status among censored (n=29)** |  |  |  |
| ≥90% healthy tissue | 2 (12.5%) | 1 (7.7%) | 3 (10.3%) |
| 75–89% healthy tissue | 0 (0.0%) | 0 (0.0%) | 0 (0.0%) |
| <75% (including 0% or missing) | 14 (87.5%) | 12 (92.3%) | 26 (89.7%) |
| **Time-to-readiness (Kaplan–Meier)** |  |  |  |
| Median time to readiness, days (95% CI bootstrap) | 12.0 (9.0–14.0) | 10.0 (7.0–22.0) | 12.0 (9.0–14.0) |
| Restricted mean time ready by $\tau=22$ days (95% CI bootstrap) | 9.85 (8.1–11.6) | 9.20 (5.7–12.5) | 9.68 (8.1–11.3) |
| **Closure method among those achieving readiness (n=59)** |  |  |  |
| Primary intention closure | 15 (33.3%) | 5 (35.7%) | 20 (33.9%) |
| Skin graft (partial thickness) | 24 (53.3%) | 7 (50.0%) | 31 (52.5%) |
| Musculocutaneous flap | 3 (6.7%) | 2 (14.3%) | 5 (8.5%) |
| Secondary intention closure | 3 (6.7%) | 0 (0.0%) | 3 (5.1%) |

### Data Source and Collection Method

Data were abstracted from the clinical records and de-identified by treating physicians at three hospitals in Gaza providing trauma care. Data extraction was performed using the KoboTool Box platform resulting in de-identified files for analysis.

### Variables

Variables extracted from clinical charts included patient demographics (age, sex); any documentation of comorbidities, especially those involving wound healing (e.g., diabetes, vascular disease); injury mechanism when recorded (e.g., gunshot, explosive projectile, crush); Wound characteristics (e.g., dimensions, location, contamination); details of initial therapies (e.g., debridement, antibiotic use); timing and wound status at interval evaluations; recorded adverse events (e.g., pain, signs of infection, bleeding, other); other surgical interventions (e.g., external fixation, flap, skin graft); and details of NPWT therapy (e.g., duration, number of dressing changes).

The primary outcome of interest was achievement of healthy granulation tissue in the wound and readiness for closure. Secondary outcomes included time to readiness for closure, method of definitive wound coverage (e.g., primary intention closure, skin graft, MC flap).

### Data Analysis

Data were analyzed using R (version 4.5.1 (2025-06-13) -- “Great Square Root” release for macOS). Descriptive statistics were performed to summarize patient characteristics. In this secondary analysis, the primary outcome, time to readiness for wound closure, was conservatively operationalized as documented definitive closure. Wound bed status alone (e.g., ≥90% healthy granulation tissue) was not considered sufficient evidence of readiness in the absence of definitive closure documentation but informed wound status at final observation (readiness for closure or right-censoring). Patients were right-censored at their last documented visit if they were lost to follow-up (LTFU) or completed observation without undergoing definitive closure. Follow-up time was measured in days from NPWT initiation to event or censoring. For LTFU patients, a study member (MH) who is a surgical specialist with experience performing war surgery made an estimate of percentage of wound coverage with healthy granulation tissue at last recorded visit from review of treating physicians’ notes and from wound photo at last visit. Kaplan–Meier methods were used to estimate the cumulative probability of readiness (1 − S(t)). Further, we estimated the restricted mean survival time (RMST), defined as the area under the survival curve up to a prespecified truncation time, τ, which was chosen as the 90th percentile of observed follow-up (τ = 22 days). RMST provides a robust, model-free summary of average time to readiness that remains interpretable even when proportional hazards’ assumptions are not met, and follow-up is incomplete. Median time to readiness and 95% confidence intervals were estimated using nonparametric bootstrap resampling (1,000).

## Ethical Considerations

This retrospective analysis of de-identified secondary clinical data was reviewed by the Yale University IRB and deemed not human subjects research (02 September 2025).

## Results

Of the 88 patients prescribed NPWT treatment, 61 (69.3%) had complete documentation of their clinical care and 27 (30.7%) were lost to follow-up (LTFU). Initial patient characteristics are summarized in Table 1. There were twice as many males (61, 69%) as females (27, 31%) in this cohort with median age of 30.1 years (IQR 18.5-41.2). Almost 90% of wounds in this cohort were on the extremities. Wounds in this cohort were complicated by infection (60, 68%), exposed bone (12, 14%), and tissue necrosis (51, 58%) (Table 2). The mechanism of injury in 49 (56%) of the wounds was result of explosion. Many wounds were likely with late presentations as more than half (46, 52%) had evidence of prior healing at initial presentation (Figure 1–3). All wounds were deemed by the treating physician to need surgical debridement and antibiotic therapy. Most cases were treated at hospital C (64, 73%) and treatment intensity was moderate requiring a median number of 2 visits (IQR 2.0 – 3.0) over a median of 7 days (IQR 2.0 – 13.2) (Table 2). Wound measurements were rarely reported and could not be included in the analysis.

**Figure 1.**
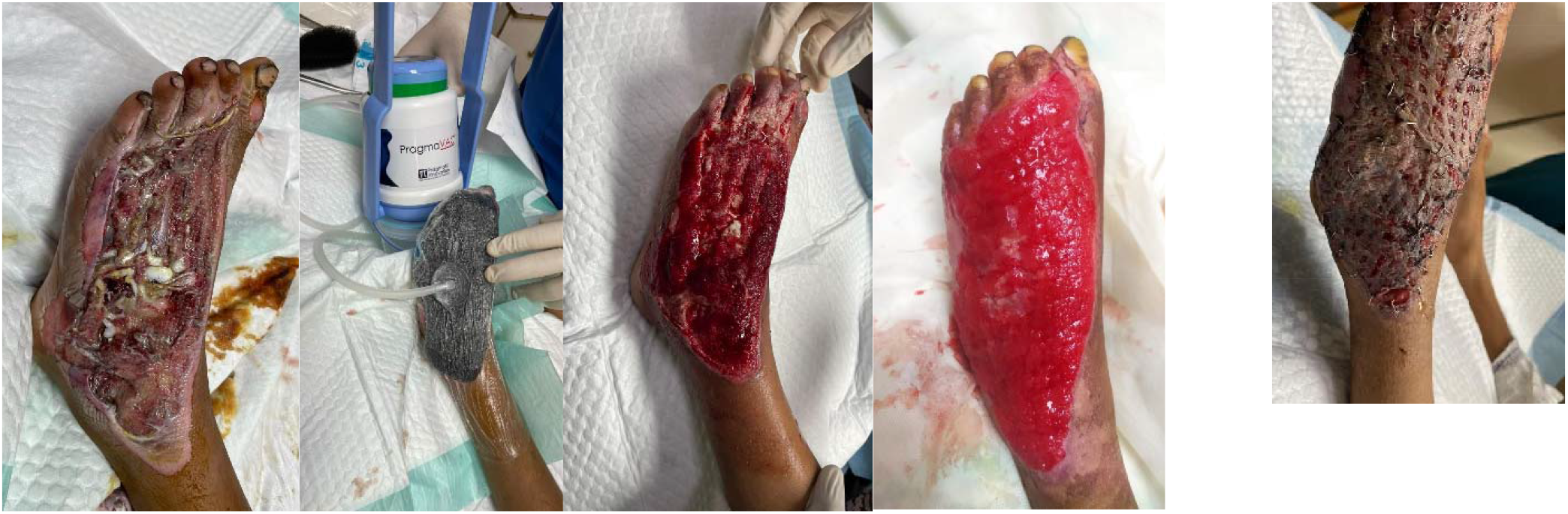
Male under 10 years old Panel A – Day 0 Initial after debridement; Panel B – NPWT sponge applied; Panel C – Day 8 Dressing Change; Panel D – Day 28; Panel E – After partial thickness skin graft

**Figure 2.**
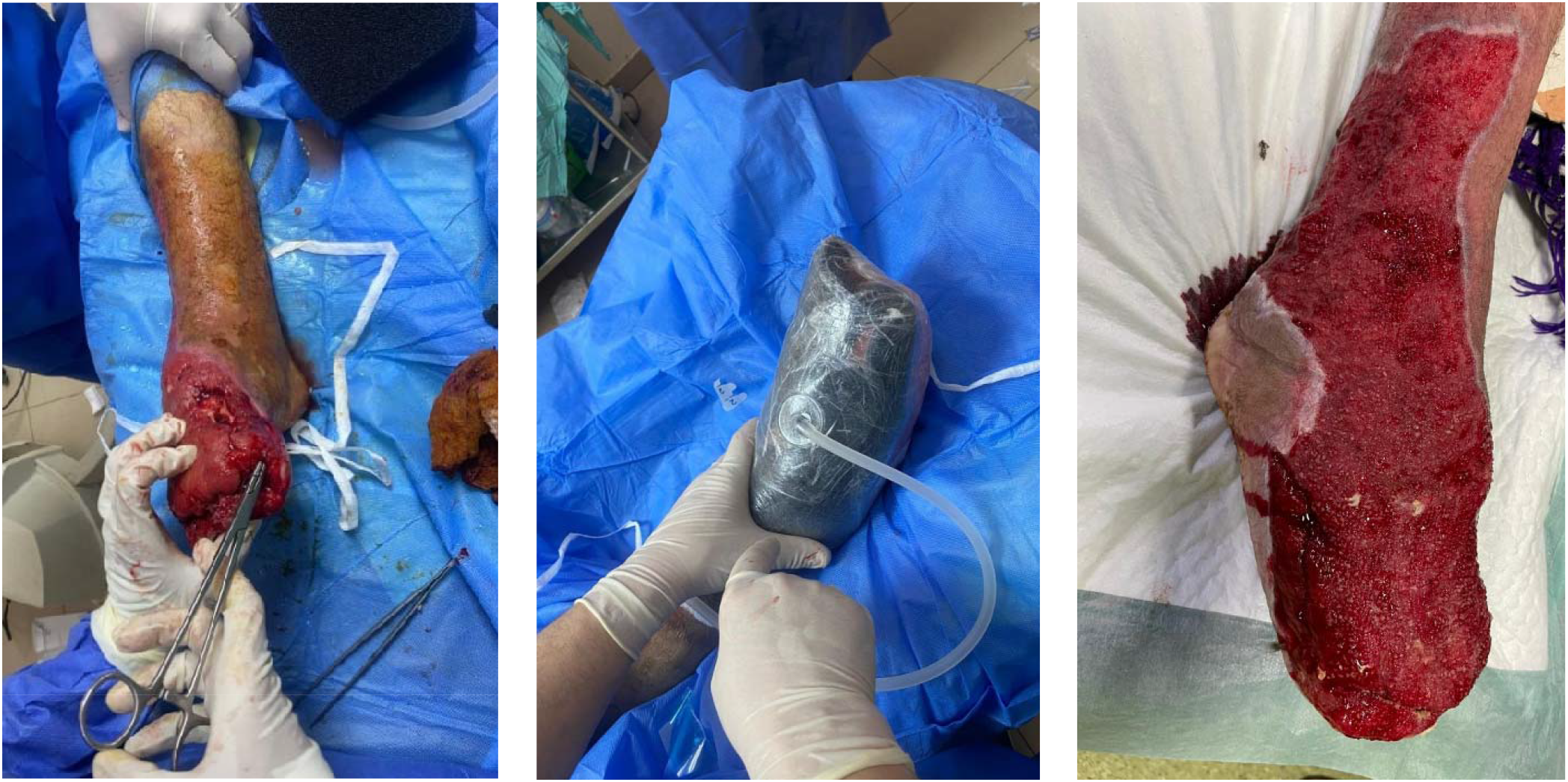
Female 21-30 years old Panel A – Day 0 Initial after debridement and partial amputation; Panel B – NPWT sponge applied Panel C – Day 5 Well vascularized wound bed ready for closure

**Figure 3.**
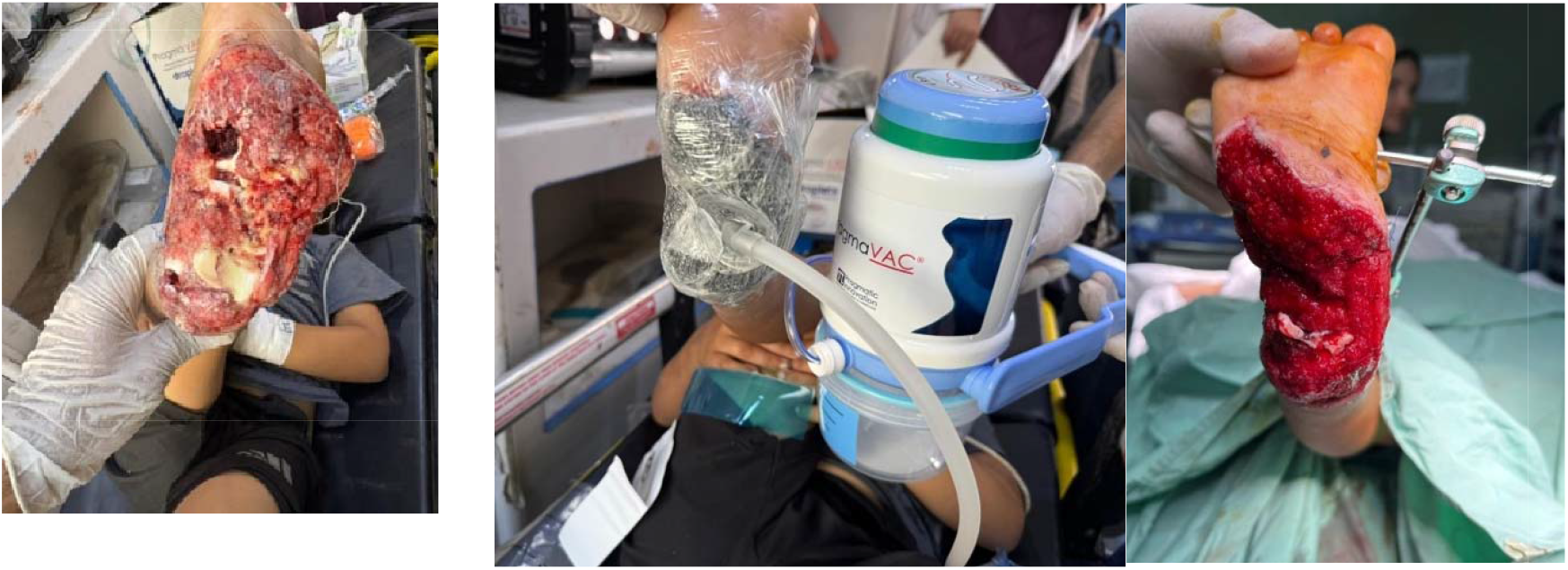
Male under 10 years old Panel A – Day 0 Initial after debridement, large soft tissue defect; Panel B – NPWT sponge attached; Panel C – Day 20 Well vascularized wound bed, no signs of infection Visit #4

Overall, 59 (67%) patients treated with the manual negative pressure wound therapy device achieved readiness for closure. Of the 29 not observed ready (right-censored), there were two treatment failures (2.3%) and remaining 27 (31%) were LTFU (Figure 4). Of those who were right-censored, 3 (10.3%) had ≥90% wound coverage with healthy granulation tissue. Median time to readiness for closure was 12 days (IQR 9.0-14.0) and adverse effects were reported in 33 cases, predominantly pain (12, 14%) and swelling (9, 10%) (Table 2). Definitive closure was achieved in 59 (67%) of cases primarily through skin grafting (31, 53%) and primary closure (20, 34%). Only 3 cases (5%) healed by secondary intention.

**Figure 4.**
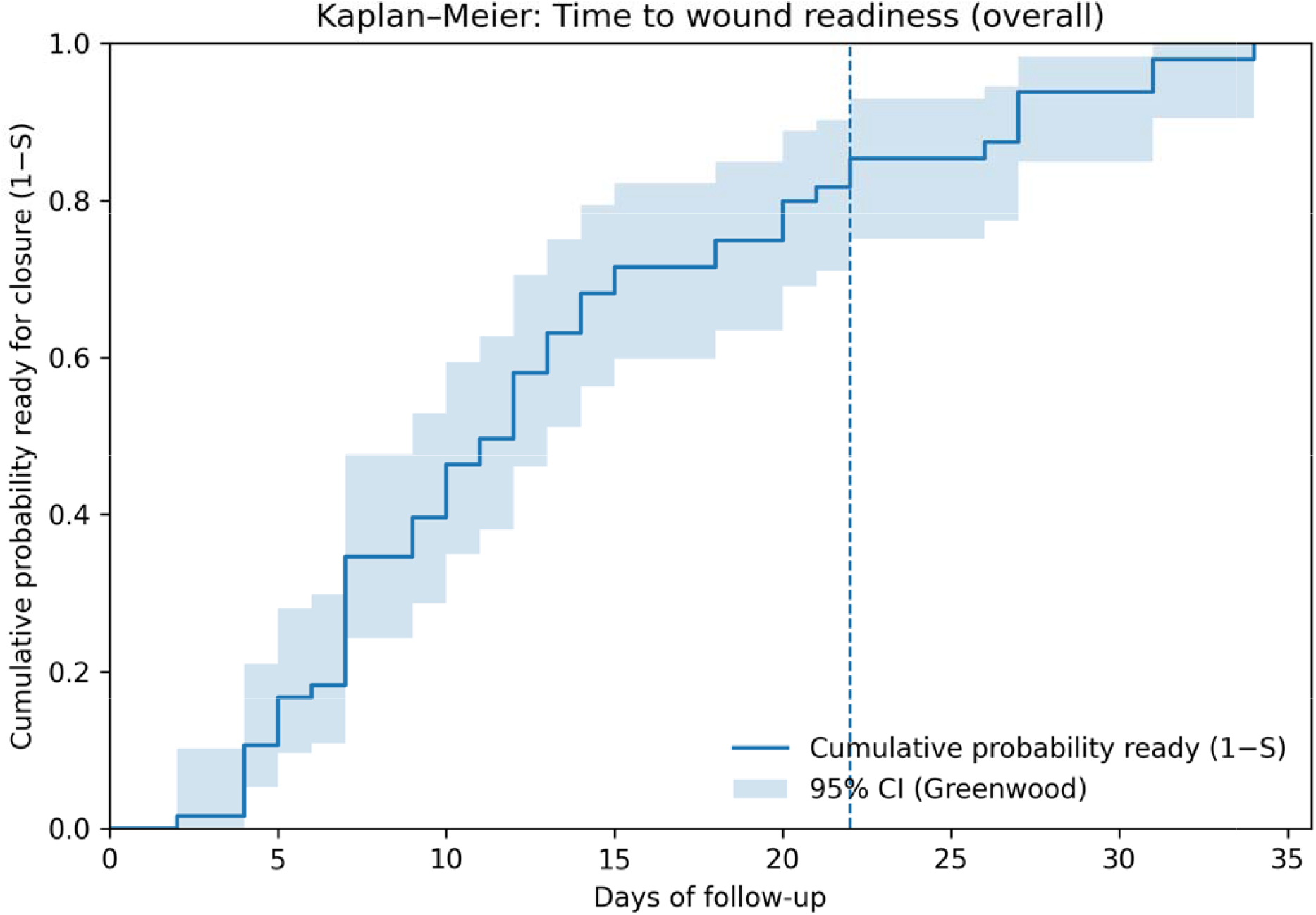
Kaplan-Meier curve demonstrating time to wound readiness foreclosure

## Discussion

The results of this retrospective cohort study indicate that the PragmaVAC NPWT device was associated with high rates of readiness for closure and few documented complications for war-related traumatic wounds in a resource-limited setting. These results build upon those of a prior prospective, randomized, controlled trial that validated the use of PragmaVAC for traumatic and atraumatic wounds.^18^ In addition to high rates of wound bed granulation and successful definitive wound coverage, these outcomes were achieved with few return visits and complications of treatment. This has important implications for the use of NPWT in war-settings where facility-based treatment is limited beyond life-saving initial trauma care, and where insecurity may limit patients’ access to medical care.

Our findings are consistent with the broader modern conflict of NPWT literature. Studies from other war settings have shown that NPWT can promote granulation, reduce dressing-change frequency, and support delayed closure or reconstruction in complex traumatic wounds. However, many of those reports involved powered NPWT systems or clinical contexts with greater access to operative care and follow-up than was available here. By contrast, the present study specifically demonstrates that a manual, electricity-independent device can be effective in a more austere environment characterized by frequent displacement, limited infrastructure, and restricted access to hospital-based care.^18,19^

While all NPWT enhance wound healing by shrinking wound defects, reducing edema, and promoting perfusion and granulation tissue formation, the manual NPWT device has several advantages for use in humanitarian settings over traditional NPWT devices including electricity independence allowing its use in settings where electricity is absent or unreliable; portability allowing patients to be safely discharged earlier and freeing up scarce hospital beds; simple design enables patients and caregivers to monitor and manipulate the device (e.g., add pressure when necessary) promoting adherence to therapy and potential for task-shifting from clinical to non-clinical personnel for portions of treatment; and small number of visits needed to achieve final outcome is beneficial in settings where insecurity limits safe patient transport and many patients are lost to follow-up. Fewer needed visits for dressing changes also help reduce demand on overburdened health facilities.

These findings extend upon the earlier experience with PragmaVAC in other conflict zones including in Syria, where it was demonstrated that the PragmaVac led to effective granulation, decreased need for dressing changes, and high patient tolerance.^18^ While that study focused on both traumatic and atraumatic wounds, our data emphasize the utility of the device specifically in managing high-energy, war-related traumatic wounds. Patient demographics and injury patterns in Gaza, including high-energy mechanism injuries like blast trauma,^20^ align with prior conflict studies, but the infrastructure challenges—such as frequent power outages, shortages in antiseptics, and increased prevalence of multidrug-resistant organisms— underscore the value of a manual, sterile, field-appropriate system.^3,20^

Clinically, the device supported key principles of wound healing: robust granulation tissue formation, infection control, and exudate management. These translated into shorter treatment durations (median 12 days) and a reduced need for repeated interventions.^18^ In this study, most cases were treated “out of the hospital” (not all patients were in home settings as many were displaced by war). In some cases, dressing changes were implemented by trained patients or caregivers outside of the clinic.

Despite the favorable device profile and efficaciousness demonstrated in our results, some barriers to effective use of NPWT devices in conflict settings like Gaza remain. Destruction of health facilities with limited access to sterile supplies, patient monitoring, and maintenance of clinical records. Patient follow-up was difficult due to both from widespread insecurity as well as frequent population displacement.^21^ The training of frontline providers in manual NPWT was challenging in contexts where surgical teams rotate frequently or are understaffed.

Beyond the device, broader determinants of wound healing must also be considered. Access to sanitation remains severely limited in many areas, especially among displaced populations.^22,23^ Clean dressing changes are compromised by poor water and compromised sanitation infrastructure.^24–26^ Nutritional deficiencies with severe food insecurity affecting 98% of households and malnutrition reaching critical levels contribute to delayed and impaired healing,^27–29^ Protein-energy malnutrition, anemia, and micronutrient deficits impair collagen synthesis and immune responses.^30^ Ideally, a holistic model of wound care should integrate nutritional assessment and supplementation into treatment pathways, though significant resource limitations exist in this setting.

## Limitations

This study presents several important limitations, primarily stemming from its retrospective nature, which introduces an inherent selection bias and limits the generalizability of the findings due to varying patient cohorts and hospital treatment practices. A significant constraint is the absence of a control group receiving standard wound care or other NPWT modalities, making it challenging to draw conclusions about the PragmaVAC device’s relative efficacy. The outcome and vital status of the 27 patients without complete documentation is unknown. Finally, patients in this study were prescribed NPWT at the discretion of their physicians with few guidelines to standardize selection for treatment.

Future studies should include prospective studies to evaluate the efficacy vs other treatment modalities (e.g., standard NPWT, traditional wet-to-dry dressings); studies of implementation should be conducted to identify facilitators and barriers to successful NPWT use in humanitarian settings, studies of patients and providers can demonstrate acceptability of NPWT device use in these settings; cost-effectiveness studies can be conducted to demonstrate the total cost per treated cost vs traditional wound care; and finally, evaluations of supply chain logistics, including local assembly or adaptation, will help determine the long-term sustainability of this technology in diverse conflict settings.

## Conclusion

Our results indicate that portable, low-cost NPWT systems such as PragmaVAC represent a clinically effective and operationally feasible innovation for traumatic wound management in conflict zones. Additional work is needed to incorporate such devices into trauma treatment protocols in humanitarian settings.

## Supporting information

STROBE checklist for observational studies

sensitivity bounds for time to readiness for closure

time to readiness for closure by sex

## Data Availability

All data produced in the present study are available upon reasonable request to the authors

## Supplemental Materials

**Figure S1.**
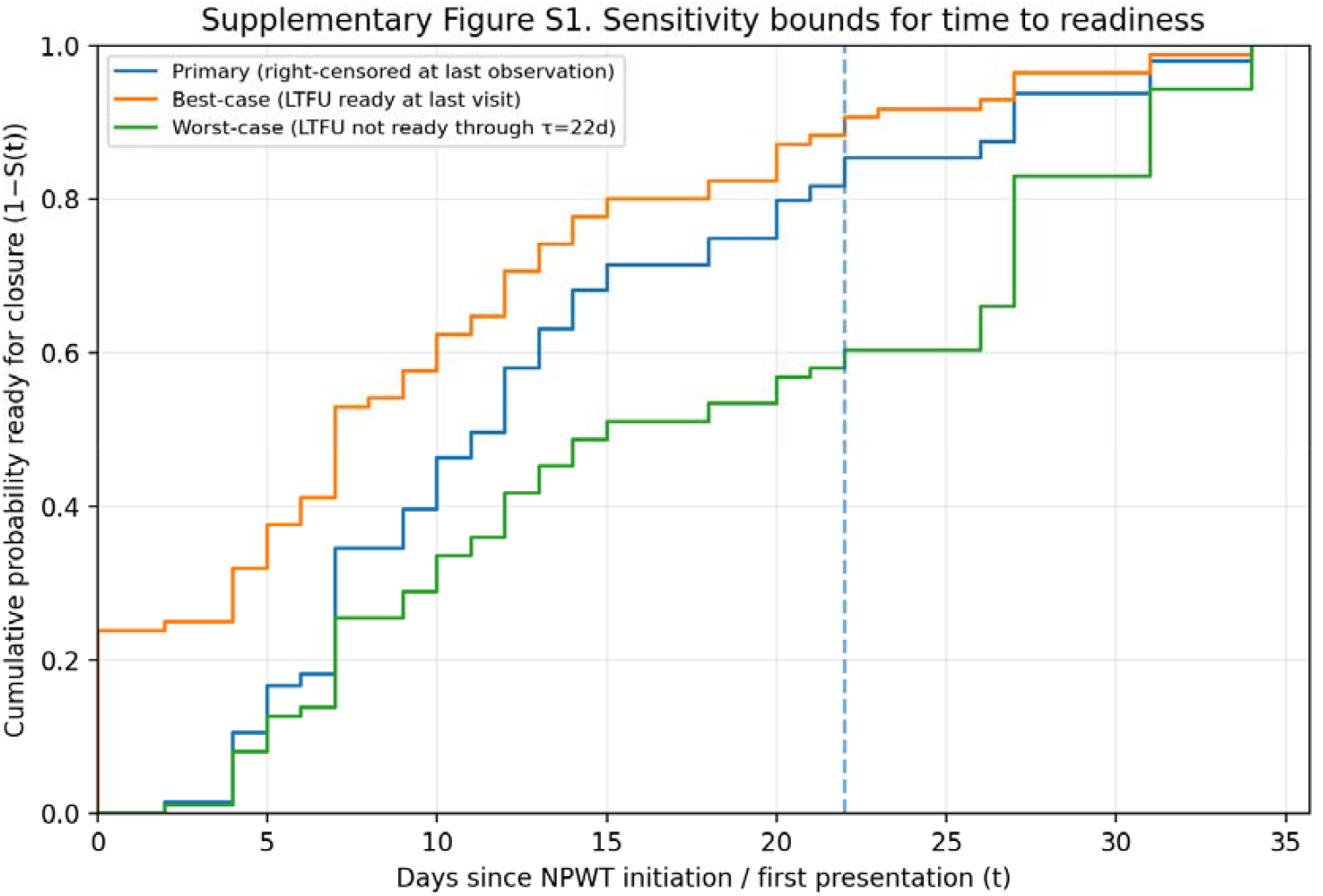
Sensitivity bounds for time to readiness.

**Figure S2.**
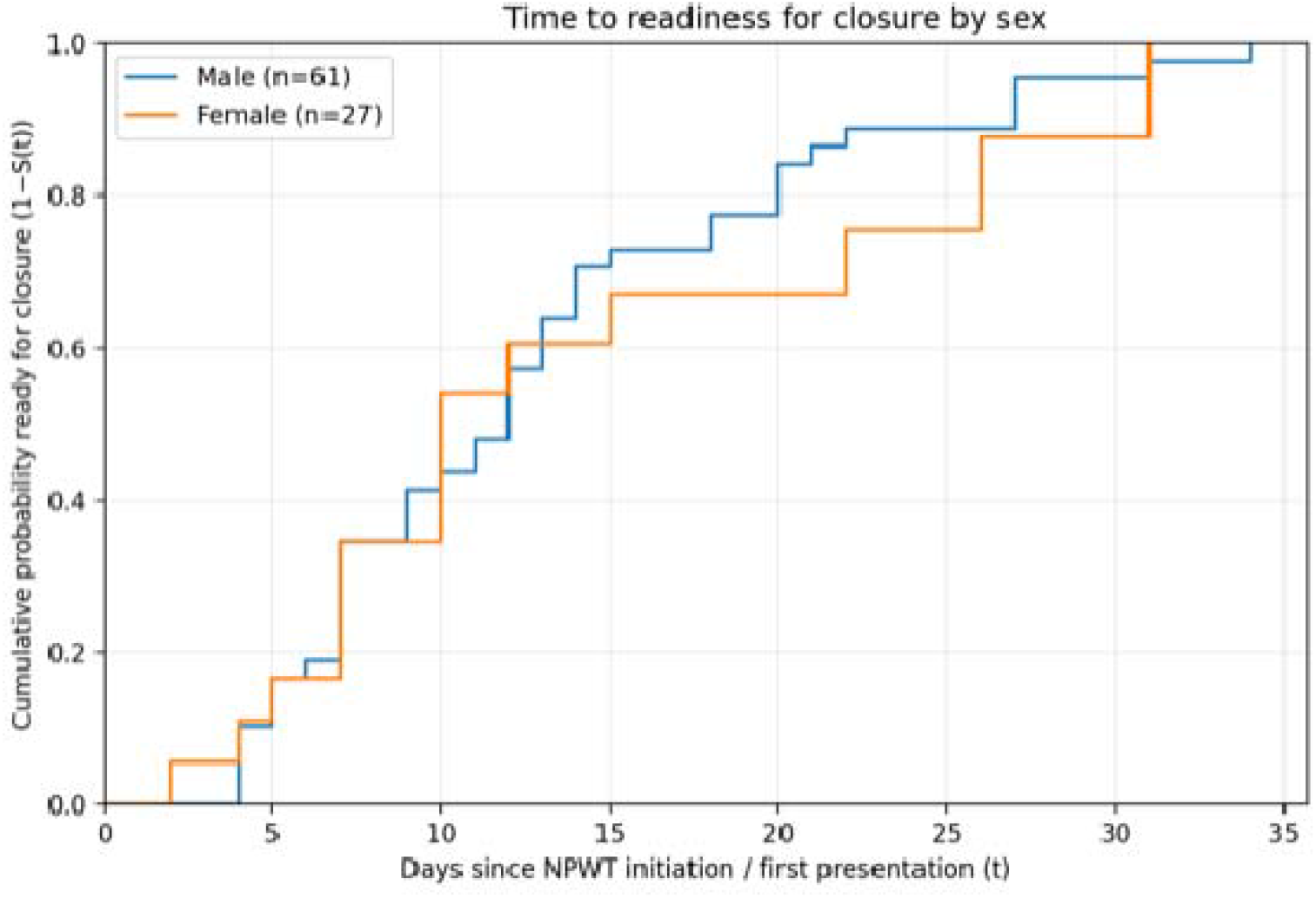
Time to readiness for closure by sex.

**Table S1.** STROBE Checklist — Cohort Study. **Manuscript:** Efficacy of the PragmaVAC Manual Negative Pressure Wound Therapy Device to Treat Acute Traumatic Wounds in a Conflict Setting: A Retrospective Cohort Study from Gaza

| Section | Item | STROBE recommendation | Reported location |
| --- | --- | --- | --- |
| Title and abstract | 1a | Indicate the study design with a commonly used term in the title or abstract | Title page; Abstract, p. 1 |
| Title and abstract | 1b | Provide an informative and balanced summary of what was done and what was found | Abstract, pp. 1–2 |
| Introduction | 2 | Explain the scientific background and rationale for the investigation | Background, pp. 2–4 |
| Introduction | 3 | State specific objectives, including any prespecified hypotheses | End of Background, p. 4 |
| Methods | 4 | Present key elements of study design early in the paper | Methods—Study Design, p. 4 |
| Methods | 5 | Describe the setting, locations, and relevant dates, including periods of recruitment, exposure, follow-up, and data collection | Methods—Study Design/Sample Population, pp. 4–5 |
| Methods | 6a | Give the eligibility criteria, and the sources and methods of selection of participants. Describe methods of follow-up | Methods—Sample Population/Data Source, p. 5 |
| Methods | 6b | For matched studies, give matching criteria and number of exposed and unexposed | Not applicable |
| Methods | 7 | Clearly define all outcomes, exposures, predictors, potential confounders, and effect modifiers | Methods—Variables/Data Analysis, pp. 5–6 |
| Methods | 8 | For each variable of interest, give sources of data and details of methods of assessment | Methods—Data Source/Collection/Variables, pp. 5–6 |
| Methods | 9 | Describe any efforts to address potential sources of bias | Methods—Data Analysis; Discussion—Limitations, pp. 6, 12–13 |
| Methods | 10 | Explain how the study size was arrived at | Methods—Sample Population, p. 5 |
| Methods | 11 | Explain how quantitative variables were handled in the analyses | Methods—Data Analysis, p. 6; Tables 1–2 |
| Methods | 12a | Describe all statistical methods, including those used to control for confounding | Methods—Data Analysis, p. 6 |
| Methods | 12b | Describe any methods used to examine subgroups and interactions | Methods—Data Analysis; Figure S2, p. 6; Supplement |
| Methods | 12c | Explain how missing data were addressed | Methods—Data Analysis; Table 2; Figure S3 |
| Methods | 12d | If applicable, explain how loss to follow-up was addressed | Methods—Data Analysis; Table 2; Figure S1; Figure S3 |
| Methods | 12e | Describe any sensitivity analyses | Methods—Data Analysis; Figure S1 |
| Results | 13a | Report numbers of individuals at each stage of study | Results; Table 2; Figure S3, pp. 6–7; Supplement |
| Results | 13b | Give reasons for non-participation at each stage | Results; Figure S3 |
| Results | 13c | Consider use of a flow diagram | Figure S3 |
| Results | 14a | Give characteristics of study participants and information on exposures and potential confounders | Results; Table 1, pp. 6–8 |
| Results | 14b | Indicate number of participants with missing data for each variable of interest | Tables 1–2; Results |
| Results | 14c | Summarize follow-up time | Table 1; Table 2; Figure 4 |
| Results | 15 | Report numbers of outcome events or summary measures over time | Results; Table 2; Figure 4; Figures S1–S2 |
| Results | 16a | Give unadjusted estimates and, if applicable, confounder-adjusted estimates and their precision | Table 2; Figure 4; Figures S1–S2 |
| Results | 16b | Report category boundaries when continuous variables were categorized | Tables 1–2 |
| Results | 16c | Translate estimates of relative risk into absolute risk for a meaningful time period | Not applicable; absolute event counts and cumulative readiness are reported |
| Results | 17 | Report other analyses done, including subgroup analyses and sensitivity analyses | Figure S1; Figure S2; Table 2 |
| Discussion | 18 | Summarize key results with reference to study objectives | Discussion, pp. 10–11 |
| Discussion | 19 | Discuss limitations, considering sources of potential bias or imprecision | Limitations, pp. 12–13 |
| Discussion | 20 | Give a cautious overall interpretation of results | Discussion/Conclusion, pp. 10–13 |
| Discussion | 21 | Discuss generalizability of the study results | Discussion/Conclusion, pp. 11–13 |
| Other information | 22 | Give the source of funding and role of funders | Acknowledgments/Funding, p. 14 |

## Acknowledgments

No funding was provided for this study.

The authors wish to acknowledge UOSSM USA and UOSSM CA who provided material support to the hospitals where patients were treated, the treating physicians and hospitals in Gaza, and the Grand Challenges grant from the Canadian government that previously supported the development of the PragmaVac NPWT device.

