## Supplementary material for "Efficacy of the PragmaVAC Manual Negative Pressure Wound Therapy Device to Treat Acute Traumatic Wounds in a Conflict Setting: A Retrospective Cohort Study from Gaza": STROBE checklist for observational studies

### STROBE Checklist — Cohort Study

| Section/Item | Recommendation | Page No. |
| --- | --- | --- |
| 1a | Indicate study design in title or abstract | 1–2 |
| 1b | Provide informative and balanced abstract | 2 |
| 2 | Scientific background and rationale | 3–5 |
| 3 | Objectives and hypotheses | 5 |
| 4 | Study design | 5 |
| 5 | Setting and dates | 5–6 |
| 6a | Eligibility criteria | 6 |
| 6b | Follow-up methods | 6–7 |
| 7 | Outcomes/exposures/confounders | 6–7 |
| 8 | Data sources and measurement | 6 |
| 9 | Bias | 7 |
| 10 | Study size | 6 |
| 11 | Quantitative variables | 6–7 |
| 12a | Statistical methods | 7 |
| 12b | Subgroup analyses | 7 |
| 12c | Missing data | 7 |
| 12d | Loss to follow-up | 7 |
| 12e | Sensitivity analyses | 7 |
| 13a | Participant flow | 8 |
| 13b | Reasons for non-participation | 8 |
| 13c | Flow diagram | Figure S2 |
| 14a | Participant characteristics | 8–9; Table 1 |
| 14b | Missing data | 8–9 |
| 14c | Follow-up time | Tables 1–2 |
| 15 | Outcome data | 9–10; Table 2 |
| 16a | Main results | Table 2 |
| 16b | Category boundaries | Table 2 |
| 17 | Other analyses | 10; Figure S1 |
| 18 | Key results | 11 |
| 19 | Limitations | 13–14 |
| 20 | Interpretation | 11–14 |
| 21 | Generalisability | 14 |
| 22 | Funding | 15 |
| 23 | Conflicts/disclosures | 15 |
