## Supplementary figures and images for "Efficacy of the PragmaVAC Manual Negative Pressure Wound Therapy Device to Treat Acute Traumatic Wounds in a Conflict Setting: A Retrospective Cohort Study from Gaza"

### sensitivity bounds for time to readiness for closure

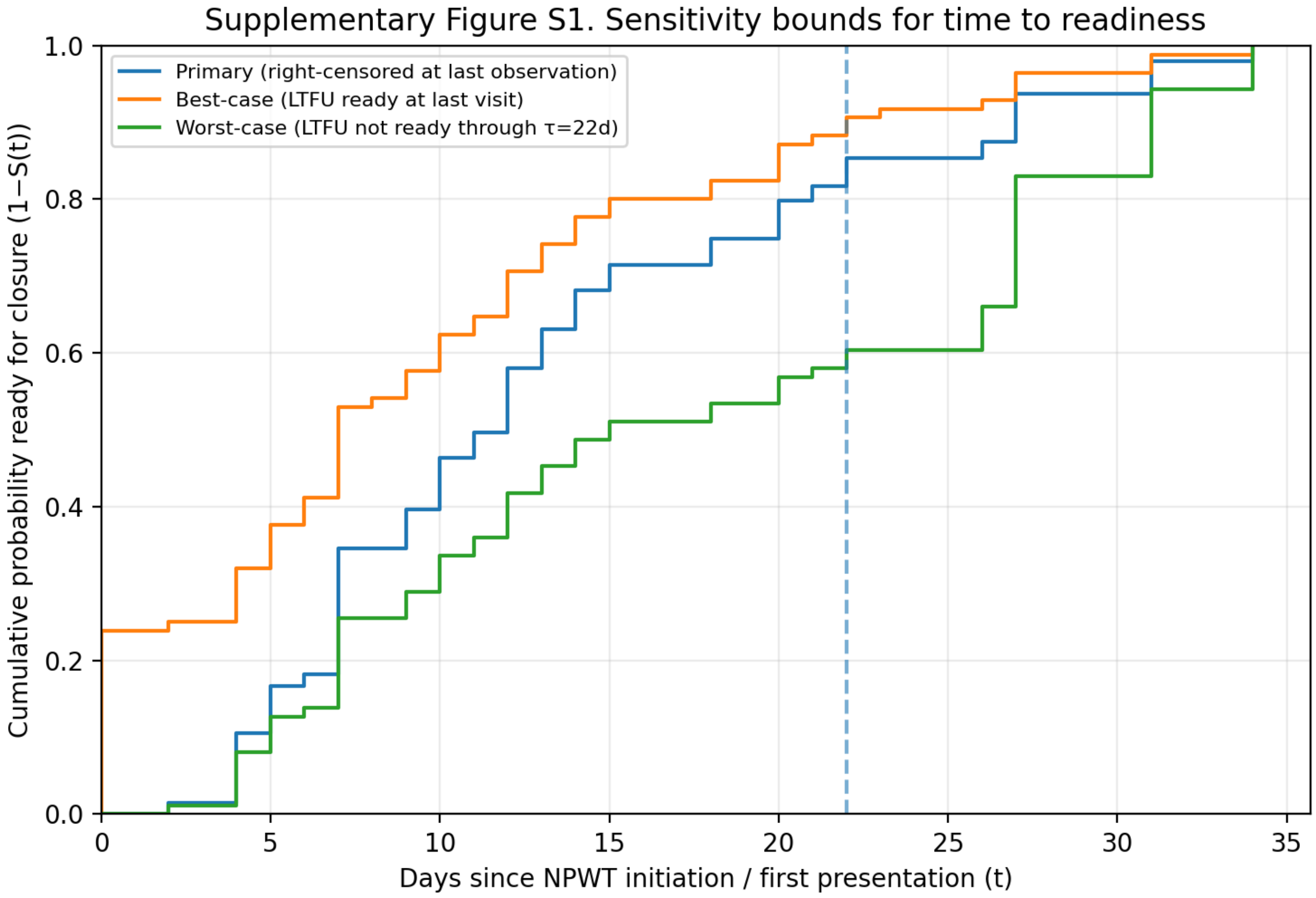

### time to readiness for closure by sex

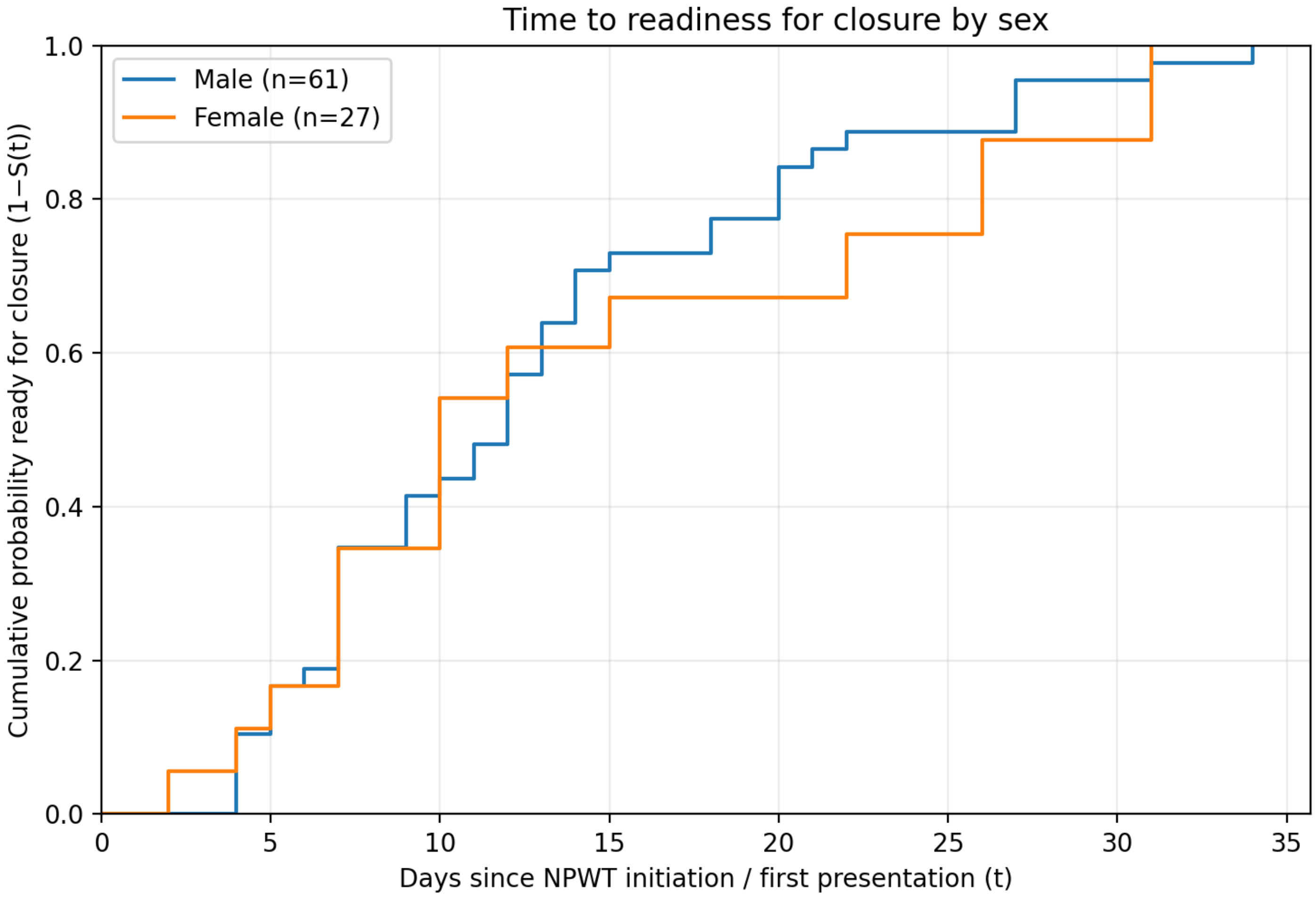
